# Effect of APOE and a polygenic risk score on incident dementia and cognitive decline in a healthy older population

**DOI:** 10.1101/2020.10.11.20210963

**Authors:** Moeen Riaz, Aamira Huq, Joanne Ryan, Suzanne G Orchard, Jane Tiller, Jessica Lockery, Robyn L Woods, Rory Wolfe, Alan E. Renton, Alison M. Goate, Robert Sebra, Eric Schadt, Amy Brodtmann, Raj C Shah, Elsdon Storey, Anne M Murray, John J McNeil, Paul Lacaze

## Abstract

**Importance:** Few studies have measured the effect of genetic factors on dementia and cognitive decline in a population of healthy older individuals followed prospectively.

**Objective:** To examine the effect of Apolipoprotein E (*APOE*) genotypes and a polygenic risk score (PRS) on incident dementia and cognitive decline in a longitudinal cohort of healthy older people.

**Design, Setting and Participants:** Post-hoc genetic analysis of a randomized clinical trial population - the ASPirin in Reducing Events in the Elderly (ASPREE) trial. At enrollment, participants had no history of diagnosed dementia, atherothrombotic cardiovascular disease, or permanent physical disability and were without cognitive impairment.

**Main Outcomes and Measures:** Dementia (adjudicated trial endpoint) and cognitive decline, defined as a >1.5 standard deviation decline in test score for either global cognition, episodic memory, language/executive function or psychomotor speed, versus baseline scores. Cumulative incidence curves for all-cause dementia and cognitive decline were calculated with mortality as a competing event, stratified by *APOE* genotypes and tertiles of a PRS based on 23 common non-*APOE* variants.

**Results:** 12,978 participants with European ancestry were included; 54.8% were female, and average age at baseline was 75 years (range 70 to 96). During a median 4.5 years of follow-up, 324 (2.5%) participants developed dementia and 503 (3.8%) died. Cumulative incidence of dementia to age 85 years was estimated to be 7.4% in all participants, 12.6% in *APOE* ε4 heterozygotes, 26.6% in ε4 homozygotes, 9.6% in the high PRS tertile, and 7.3% in the low PRS tertile. *APOE* ε4 heterozygosity/homozygosity was associated with a 2.5/6.3-fold increased risk of dementia and a 1.4/1.8-fold increased risk of cognitive decline, versus ε3/ε3 (P<0.001 for both). A high PRS (top tertile) was associated with a 1.4-fold increase risk of dementia, versus the low tertile (CI 1.04-1.76, P=0.02), but was not associated with cognitive decline risk (CI 0.96-1.22, P = 0.18).

**Conclusions and Relevance:** Incidence of dementia among healthy older individuals is low across all genotypes; however, *APOE* ε4 and high PRS increase relative risk. *APOE* ε4 is associated with cognitive decline, but PRS is not.

**KEY POINTS:** 

**Question:** How do genetic factors influence the risk of dementia and cognitive decline among healthy older individuals?

**Findings:** We studied cumulative incidence of dementia and cognitive decline in 12,978 healthy older individuals without cardiovascular disease or cognitive impairment at enrollment, stratified by *APOE* genotype and a polygenic risk score (PRS). *APOE* ε4 and PRS increased the relative risk of dementia, but cumulative incidence was low across all genotypes. *APOE* genotypes were associated with cognitive decline, but PRS was not.

**Meaning:** Incidence of dementia is low among healthy older individuals; however, genetic factors still increase relative risk.

## INTRODUCTION

Few studies have measured the effect of Apolipoprotein E (*APOE*) genotypes and polygenic risk scores (PRS) on incident dementia and cognitive decline in healthy older people. The ASPREE (ASPirin in Reducing Events in the Elderly) cohort offers the opportunity to measure these effects, as recruited participants had no history of cardiovascular disease, dementia or significant physical disability at enrollment. The ASPREE study was a randomised, placebo-controlled trial to determine whether daily low dose aspirin increased survival, free of dementia or persistent physical disability, in 19,114 healthy community-dwelling older people(1). In 2018, ASPREE reported that over an average 4.5 year follow-up, aspirin did not prolong disability-free survival(2-4) or reduce the risk of dementia or cognitive decline(5).

The *APOE* gene is the strongest genetic determinant of all-cause dementia, especially Alzheimer’s disease (AD), with the ε4 allele elevating risk and accelerating age of onset(6-9). The ε4 allele is also associated with cognitive impairment (dysfunction in episodic memory, processing speed, executive function or global cognition) in people without a dementia diagnosis(10-13). Beyond *APOE*, common disease-associated variants identified from genome-wide association studies (GWAS)(14-18) also modify dementia risk, and can be used to calculate a polygenic risk score (PRS)(9, 19-24). Individually, these common genetic variants have low effect sizes, yet when combined into a PRS can enable risk-stratification for dementia indications beyond *APOE* genotype. There is varying evidence for whether a PRS for dementia can also predict cognitive decline(25-28). Incorporating both *APOE* genotypes and PRS, alongside conventional risk factors, may enable more accurate risk prediction(29, 30). This may aid development of therapeutic strategies or prevention, and advance our understanding of the genetic differences between (diagnosed) dementia, and cognitive decline.

The predictive performance of PRSs for dementia requires further investigation in well-characterised prospective studies. Predictive performance can be influenced by factors such as ethnicity, age, study recruitment criteria, clinical diagnostic criteria, neuropsychological assessments used, genotyping platform, and genetic variants included(6-9, 19-24, 29-31). More studies of cognitively healthy elderly individuals followed prospectively are required to assess variability and predictive accuracy. Here, we report the effects of *APOE* and PRS on incident dementia and cognitive decline among 12,978 ASPREE participants, where dementia was an exclusionary criterion at entry and adjudicated as a primary trial endpoint.

## METHODS

### Study population

Consistent with the ASPREE inclusion criteria(1), participants had no previous history or current diagnosis of atherothrombotic cardiovascular disease, dementia, loss of independence with basic activities of daily living, or life-threatening illness. Participants passed a global cognition screen at enrollment (>77 on the Modified Mini-Mental State (3MS) Examination). Informed consent for genetic analysis was obtained, with ethical approval from the Alfred Hospital Human Research Ethics Committee (390/15) and site-specific Institutional Review Boards (US).

### Incident Dementia Diagnosis

After standardized cognition and functional measures, participants reporting memory or cognitive problems were assessed by specialists or prescribed dementia medication (Australia). Following identification of dementia triggers (3MS<78 or a drop of >10.15 points from the participant’s baseline 3MS score, accounting for age and education), additional assessments were conducted, with brain imaging and laboratory analyses collected for adjudication. Each dementia trigger case was reviewed according to the ASPREE protocol for clinical adjudication(4, 5) by an adjudication committee consisting of geriatricians, neurologists and neuropsychologists. Dementia was diagnosed using Diagnostic and Statistical Manual of Mental Disorders, fourth edition criteria. Diagnosis date was recorded as date of trigger.

### Cognitive Decline

The ASPREE cognitive battery included the 3MS for general cognition, the Hopkins Verbal Learning Test-Revised (HVLT-R) for episodic memory, the single letter Controlled Oral World Association Test (COWAT) for language and executive function, and the Symbol Digit Modalities Test (SDMT) to measure psychomotor speed. Accredited professionals administered assessments at baseline and year 1, followed biennially during follow-up. As reported previously(5), cognitive decline in participants without a dementia diagnosis was defined as a 1.5 standard deviation decline in 3MS/HVLT-R/SDMT/COWAT compared with baseline scores, sustained over ≥2 timepoints.

### Genotyping and variant analysis

14,052 samples were genotyped using the Axiom 2.0 Precision Medicine Diversity Research Array following standard protocols. 12,978 samples passed quality control (12,343 Australian, 635 US) based on sex, relatedness and ancestry (Non-Finnish Europeans). To estimate population structure, we performed principal component analysis using the 1000 Genomes reference population (Figure S1)(32, 33). Imputation was performed using the haplotype reference consortium European panel(34). Post-imputation quality control removed variants r2<0.3. *APOE* genotype was measured using two directly genotyped variants (rs7412, rs429358) extracted using plink v1.9(35).

### Polygenic risk score

PRS was calculated using 23 common variants (15 genotyped, 8 imputed) associated with AD at genome-wide significance, that affect risk independently of *APOE*(17, 24, 36). PRS calculations, using plink v1.9(35), were based on dosage (0,1,2) of SNP effect allele reported from GWAS, multiplied by effect sizes, followed by the sum of products to generate a PRS per participant (Table S1). PRS distribution was divided into low/middle/high tertiles; with mean values of; low −0.56 (range −1.43 to −0.34), middle −0.20 (−0.34 to −0.06), high 0.16 (−0.06 to 1.86)(Figure S2).

### Statistical analysis

To determine whether *APOE* genotype frequencies were under selective pressure due to age and/or trial inclusion/exclusion criteria, we performed Hardy-Weinberg equilibrium (HWE) testing. This compared observed genotype frequencies with those expected in a population under no selective pressure, using chi-squared tests. We examined the cumulative incidence of dementia (CID) and cognitive decline, stratified by *APOE* genotype and PRS tertiles. We used ε3 homozygotes as a reference group for *APOE*-stratified analysis, and the low-risk tertile for PRS-stratified analysis. Consistent with other studies(9, 24) we combined *APOE* ε3/ε4:ε2/ε4 into a single group, and ε2/ε2:ε2/ε3 into a single group.

We estimated cumulative incidence of all-cause dementia and cognitive decline during an average of 4.5 years of follow-up, using the Cumulative Incidence Function (CIF) of the etm package(37, 38) in R version 3.6.0(39). Data were censored by date of dementia diagnosis, cognitive decline, last contact or death. The age on censored date was used as a time scale in CIF model. Cumulative incidence was calculated up to 95 years, then stratified by *APOE* genotype and PRS tertiles. Dementia and cognitive decline between PRS tertiles was compared for the whole cohort and further stratified by *APOE* genotypes. The dementia and cognitive decline models were estimated independently.

We used the Fine and Gray (F&G) method of accounting for competing risk of death, and Cox proportional hazard regression model to calculate dementia hazard ratio of both models, for *APOE*, PRS and their interaction, adjusted for age at enrollment (continuous, allowing a quadratic function) and sex(9, 37). We used age on censored date as a time scale in both F&G and Cox models. Hazard ratios for cognitive decline were measured the same way. To test association of *APOE* genotypes and PRS with cohort characteristics, we used a multivariable regression model with variables; age, sex, follow-up time, education, alcohol use, smoking, diabetes mellitus, hypertension, depression (Center for Epidemiological Studies-Depression-10 scale), family history of dementia (father/mother/sibling), body mass index, blood pressure, cholesterol, triglycerides. Bonferroni multiple test correction at P=0.002 significance was applied (0.05/17=0.002).

## RESULTS

Characteristics of the 12,978 genotyped participants are shown in Table 1. Overall, 54.8% were female, 47% had educational attainment <12 years, 3% were current smokers, and 25% reported a family history of dementia at enrollment.

**Table 1:** Characteristics of the ASPREE cohort stratified by *APOE* genotypes and tertiles of a PRS.

|  | All | ε3/ε3 | ε3/ε4 | ε2/ε2 | ε2/ε3 | ε2/ε4 | ε4/ε4 | Low PRS<br>tertile | Middle PRS<br>tertile | High PRS<br>tertile |
| --- | --- | --- | --- | --- | --- | --- | --- | --- | --- | --- |
| **Total** | 12978 | 7800(60.0%) | 2665(20.5%) | 68(0.5%) | 1784(14.0%) | 461(3.5%) | 200(1.5%) | 4326(33.3%) | 4330(33.4%) | 4322(33.3%) |
| **Demographics** |  |  |  |  |  |  |  |  |  |  |
| **Age, Years** | 75.05(4.2) | 75.12(4.3) | 74.66(3.9) | 74.97(3.9) | 75.41(4.4) | 75.28(4.4) | 73.91(3.4)* | 75.15(4.3) | 75.04(4.2) | 74.95(4.1) |
| **Follow up**<br>**time, Years** | 4.53(1.3) | 4.54(1.3) | 4.50(1.3) | 4.63(1.3) | 4.54(1.3) | 4.63(1.2) | 4.46(1.3) | 4.51(1.3) | 4.52(1.3) | 4.50(1.3) |
| **Sex** |  |  |  |  |  |  |  |  |  |  |
| **Females** | 7120(55.0%) | 4407(56.0) | 1413(53.0%) | 40(58.8%) | 916(51.3%) | 245(53.0%) | 99(49.5%) | 2385(55.0%) | 2371(54.7%) | 2364(54.6%) |
| **Males** | 5858(45.0%) | 3393(44.0) | 1252(47.0%) | 28(41.2%) | 868(48.7%) | 216(47%) | 101(50.5%) | 1941(45.0%) | 1959(45.3%) | 1958(45.4%) |
| **Education** |  |  |  |  |  |  |  |  |  |  |
| **< 12 years** | 6136(47.2%) | 3640(46.6%) | 1249(46.9%) | 27(39.7%) | 886(49.6%) | 230(49.9%) | 104(52.0%) | 2030(46.9%) | 2061(47.6%) | 2045(47.4%) |
| **12-15 years** | 3483(26.8%) | 2118(27.0%) | 707(26.5%) | 18(26.4%) | 483(27.10%) | 114(24.7%) | 43(21.5%) | 1137(26.3%) | 1178(27.2%) | 1168(27.0%) |
| **16+ years** | 3359(26.0%) | 2042(26.4%) | 709(26.6) | 23(33.9%) | 415(23.3%) | 117(25.4%) | 53(26.5%) | 1159(26.8%) | 1091(25.1%) | 1109(25.6%) |
| **Alcohol Use** |  |  |  |  |  |  |  |  |  |  |
| **Current** | 10353(79.8%) | 6254(80.2%) | 2108(79.0%) | 55(81.0%) | 1421(79.6%) | 355(77.0%) | 160(80%) | 3438(79.4%) | 3473(80.2%) | 3442(79.7%) |
| **Former** | 620(4.8%) | 372(4.8%) | 129(4.8%) | 0(0%) | 89(5.0%) | 21(4.6%) | 9(4.5%) | 210(4.9%) | 209(4.8%) | 201(4.6%) |
| **Never** | 2005(15.4%) | 1174(15.0%) | 428(16.2%) | 13(19.0%) | 274(15.4%) | 85(18.4%) | 31(15.5%) | 678(15.7%) | 648(15.0%) | 679(15.7%) |
| **Smoking status** |  |  |  |  |  |  |  |  |  |  |
| **Current** | 394(3.0%) | 244(3.1%) | 83(3.1%) | 2(3.0%) | 44(2.5%) | 11(2.4%) | 10(5.0%) | 115(2.6%) | 137(3.2%) | 142(3.2%) |
| **Former** | 5339(41.2%) | 3165(40.6%) | 1111(41.7%) | 29(42.6%) | 749(42.0%) | 195(42.3%) | 90(45.0%) | 1834(42.4%) | 1738(40.1%) | 1767(41.0%) |
| **Never** | 7245(55.8%) | 4391(56.3%) | 1471(55.2%) | 37(54.4%) | 991(55.5%) | 255(55.3%) | 100(50.0%) | 2377(55.0%) | 2455(56.7%) | 2413(55.8%) |
| **Medical History** |  |  |  |  |  |  |  |  |  |  |
| **Diabetes** | 1205(9.2%) | 747(9.5%) | 223(8.3%) | 7(10.2%) | 173(9.7%) | 41(8.8%) | 15(8.0%) | 396(9.1%) | 399(9.2%) | 410(9.4%) |
| **Hypertension** | 9553 (73.6%) | 5773(74.0%) | 1927(31.0%) | 48(70.5%) | 1330(74.5%) | 341(7.4%) | 137(68.5%) | 3172(73.3%) | 3202(73.9%) | 3178(73.5%) |
| **Depression** | 1177 (9.0%) | 702(9.8%) | 232(8.7%) | 6(8.8%) | 172(9.6%)* | 42(9.1%) | 23(11.5%)* | 396(9.1%) | 390(9.0%) | 391(9.0%) |
| **Dementia**<br>**Family**<br>**History+** | 3293(25.4%) | 1824(23.4%) | 846(31.7%)** | 11(16.1%) | 399(22.4%) | 118(25.6%) | 95(47.5%)** | 1079(25.0%) | 1053(24.3%) | 1161(26.8%)* |
| **Physical Examination** |  |  |  |  |  |  |  |  |  |  |
| **Body Mass**<br>**Index** | 27.97(4.5) | 28.08(4.6) | 27.62(4.4)* | 29.60(5.4)* | 28.09(4.4) | 27.74(4.3) | 27.66(5.0)* | 27.98(4.6) | 28.04(4.6) | 27.90(4.5) |
| **Systolic Blood**<br>**Pressure (mm**<br>**Hg)** | 139.46(16.3) | 139.37(16.2) | 139.40(16.1) | 139.44(16.2) | 139.95(16.4) | 140.05(17.0) | 138.1(15.3) | 139.50(16.2) | 139.62(16.4) | 139.26(16.2) |
| **Diastolic blood Pressure (mm Hg)** | 77.17(9.9) | 77.80(9.8) | 77.19(10.0) | 77.69(8.8) | 77.40(10.2) | 77.79(10.0) | 76.49(10.0) | 76.88(9.9) | 77.51(9.9) | 77.12(10.0) |
| **Blood Biochemistry** |  |  |  |  |  |  |  |  |  |  |
| **TC (mmol/L)** | 5.26(0.97) | 5.28(0.96) | 5.34(1.0) | 4.65(0.90)* | 5.07(0.91)* | 5.24(0.95) | 5.36(1.0) | 5.27(0.98) | 5.27(0.96) | 5.25(0.96) |
| **LDL (mmol/L)** | 3.07(0.86) | 3.11(0.86) | 3.17(0.89) | 2.31(0.73) | 2.81(0.76) | 2.99(0.84) | 3.22(0.91) | 3.08(0.87) | 3.08(0.86) | 3.07(0.86) |
| **HDL (mmol/L)** | 1.58(0.45) | 1.58(0.45) | 1.57(0.46)* | 1.63(0.41) | 1.61(0.47)* | 1.60(0.46) | 1.50(0.46) | 1.59(0.45) | 1.58(0.46) | 1.57(0.45) |
| **TG(mmol/L)** | 1.31(0.65) | 1.29(0.60) | 1.30(0.69) | 1.54(0.62)* | 1.38(0.71)* | 1.39(0.76) | 1.42(0.83) | 1.31(0.65) | 1.31(0.63) | 1.32(0.66) |
| **Disease/Trait Incidence** |  |  |  |  |  |  |  |  |  |  |
| **Dementia** | 324 (2.5%) | 147(1.8%) | 125(4.7%) | 0(0%) | 25(1.4) | 9(1.9) | 18(9.0%) | 99(2.2%) | 96(2.2%) | 129(2.9%) |
| **Cognitive Decline<sup>\$</sup>** | 1598 (12.6%) | 896(11.1%) | 385(14.4%) | 8(1.4%) | 208(11.6%) | 67(6.9%) | 34(17.0%) | 518(12.0%) | 534(12.3%) | 546(12.6%) |
| APOE: Apolipoprotein E, PRS: Polygenic risk score, LDL: low density lipoprotein, HDL: high density lipoprotein, TC: total cholesterol, TG: triglycerides, mmol/L: Millimoles per litre, mm Hg: millimetre of mercury, The numbers reported as either n(%) or mean(SD), + sign represents family history of dementia in either Father, mother or sibling. * Denotes p value < 0.05, as measured using a regression model with *APOE* ε3/ε3 and low PRS tertile as reference groups. ** Denotes associations that remained significant after Bonferroni correction for multiple testing. \$Denotes the number of samples with cognitive decline out of total 12,978 genotyped samples excluding 324 dementia cases and 326 participants with missing data. | | | | | | | | | | |

Associations of cohort characteristics with *APOE* genotypes and PRS tertiles are also shown in Table 1. The only associations to survive multiple testing correction was family history of dementia in ε4 heterozygotes/homozygotes, with no cohort characteristics differing between PRS tertiles (Table 1).

We found that *APOE* genotype frequencies had deviated from HWE (chi-square=38, P=<0.001)(Table S2), with fewer ε3/ε4 heterozygotes (N=2665 observed, N=2723 expected) and fewer ε4/ε4 homozygotes (N=200 observed, N=239 expected) than expected under HWE.

During mean 4.5 years of follow-up (interquartile range 2.1 to 5.7 years, 2779 person-years), we observed 324 (2.5%) incident all-cause dementia cases and 503 (3.8%) deaths (Table 2). For cumulative incidence of dementia (CID), we describe results up to age 85 years, representing an approximate centre point between lower and upper age ranges of the ASPREE population at baseline (70 to 96 years). CID in ASPREE was estimated at 7.4% (CI 6.5 to 8.5).

**Table 2:**
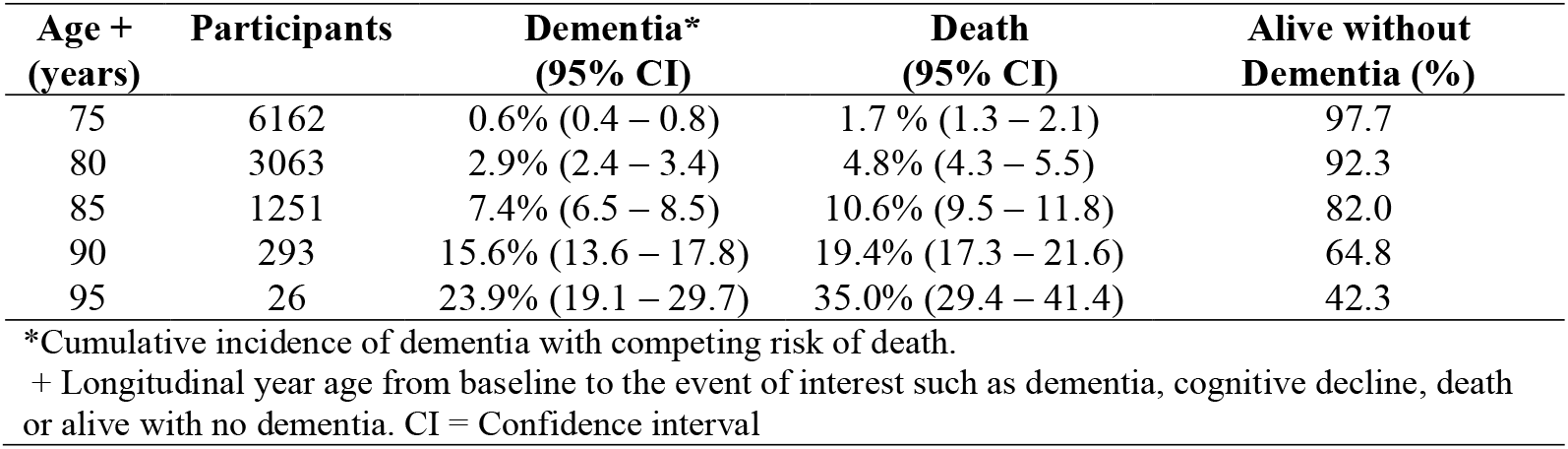
Cumulative incidence of dementia and death in ASPREE.

| **Age +**<br>**(years)** | **Participants** | **Dementia\***<br>**(95% CI)** | **Death**<br>**(95% CI)** | **Alive without**<br>**Dementia (%)** |
| --- | --- | --- | --- | --- |
| 75 | 6162 | 0.6% (0.4 – 0.8) | 1.7 % (1.3 – 2.1) | 97.7 |
| 80 | 3063 | 2.9% (2.4 – 3.4) | 4.8% (4.3 – 5.5) | 92.3 |
| 85 | 1251 | 7.4% (6.5 – 8.5) | 10.6% (9.5 – 11.8) | 82.0 |
| 90 | 293 | 15.6% (13.6 – 17.8) | 19.4% (17.3 – 21.6) | 64.8 |
| 95 | 26 | 23.9% (19.1 – 29.7) | 35.0% (29.4 – 41.4) | 42.3 |

\*Cumulative incidence of dementia with competing risk of death.
+ Longitudinal year age from baseline to the event of interest such as dementia, cognitive decline, death or alive with no dementia. CI = Confidence interval

In *APOE* genotype-stratified analysis of CID, after adjusting for covariates and death as a competing event, ε4/ε4 genotype was significantly associated with dementia risk (HR 6.38 [CI 3.8-10.7] P=2.0×10^−12^) compared with ε3/ε3 (Table 3a). Individuals with ε3/ε4 heterozygosity were also at higher risk of dementia (HR 2.51 [CI 1.9-3.1], P=1.5×10^−14^) compared with ε3/ε3. CID was 26.6% (CI 16.2-42.0) for ε4/ε4 homozygotes, 12.6% (CI 10.2-15.5) for ε3/ε4 heterozygotes, 5.9% (CI 4.8- 7.2) for the common ε3/ε3 genotype group, and 4.0% (CI 2.4-6.5) for the lower-risk ε2/ε2:ε2/ε3 group (Figure 1a). For all *APOE* genotype-stratified results see Table S3.

**Table 3:** Cox proportional hazard ratio and risk regression models for dementia and cognitive decline risk in the ASPREE cohort.

| **a) Dementia** | **§No competing risk adjustment** |  | **^Adjusting for competing risk** |  |
| --- | --- | --- | --- | --- |
| Variables | HR (95% CI) | P value | HR (95% CI) | P value |
| Age, years | 1.16 (1.13 – 1.18) | 0.002 | 1.25 (1.14 – 1.37) | <0.0001* |
| Sex (female) | 0.76 (0.61 – 0.94) | 0.01 | 0.77 (0.62 – 0.96) | 0.02 |
| *APOE* ε3ε3 | Reference | - | Reference | - |
| *APOE* ε2ε2:ε2ε3 | 0.67 (0.43-1.02) | 0.06 | 0.65 (0.43 -1.00) | 0.05 |
| *APOE* ε3ε4:ε2ε4 | 2.50 (1.97-3.16) | <0.0001<sup>+</sup> | 2.51 (1.98 - 3.17) | <0.0001<sup>+</sup> |
| *APOE* ε4ε4 | 6.32 (3.86-10.34) | <0.0001<sup>+</sup> | 6.38 (3.81-10.71) | <0.0001<sup>+</sup> |
| Low PRS tertile | Reference | - | Reference | - |
| Middle PRS tertile | 1.00 (0.75-1.32) | 0.98 | 1.00 (0.76 -1.33) | 0.95 |
| High PRS tertile | 1.36 (1.04-1.76) | 0.02 | 1.36 (1.04-1.77) | 0.02 |
| **b) Cognitive decline** | **§No competing risk adjustment** |  | **^Adjusting for competing risk** |  |
| Variables | HR (95% CI) | P value | HR (95% CI) | P value |
| Age (years) | 1.06 (1.05 – 1.07) | 0.002 | 1.05 (1.04 – 1.06) | <0.0001* |
| Sex (female) | 0.87(0.79 – 0.96) | 0.007 | 0.89 (0.81 – 0.98) | 0.02 |
| *APOE* ε3ε3 | Reference | - | Reference | - |
| *APOE* ε2ε2:ε2ε3 | 0.67(0.43-1.02) | 0.06 | 0.99(0.85-1.15) | 0.96 |
| *APOE* ε3ε4:ε2ε4 | 1.35(1.20-1.51) | <0.0001<sup>+</sup> | 1.35(1.20 -1.51) | <0.0001<sup>+</sup> |
| *APOE* ε4ε4 | 1.75(1.24-2.46) | 0.001 | 1.74(1.22 -2.47) | 0.001 |
| Low PRS tertile | Reference | - | Reference | - |
| Middle PRS tertile | 1.02(0.91-1.16) | 0.64 | 1.03(0.91-1.16) | 0.63 |
| High PRS tertile | 1.08(0.96-1.22) | 0.18 | 1.10(0.95-1.21) | 0.22 |
| § COX proportional hazard models<br>^ Risk regression model by Fine and Gray Method<br>CI: Confidence interval, HR: Hazard ratio, PRS: Polygenic risk score<br>*Denotes p values = $1.0 \times 10^{-16}$, + denotes $P < 16.3 \times 10^{-12}$ | | | | |

**Figure 1.**
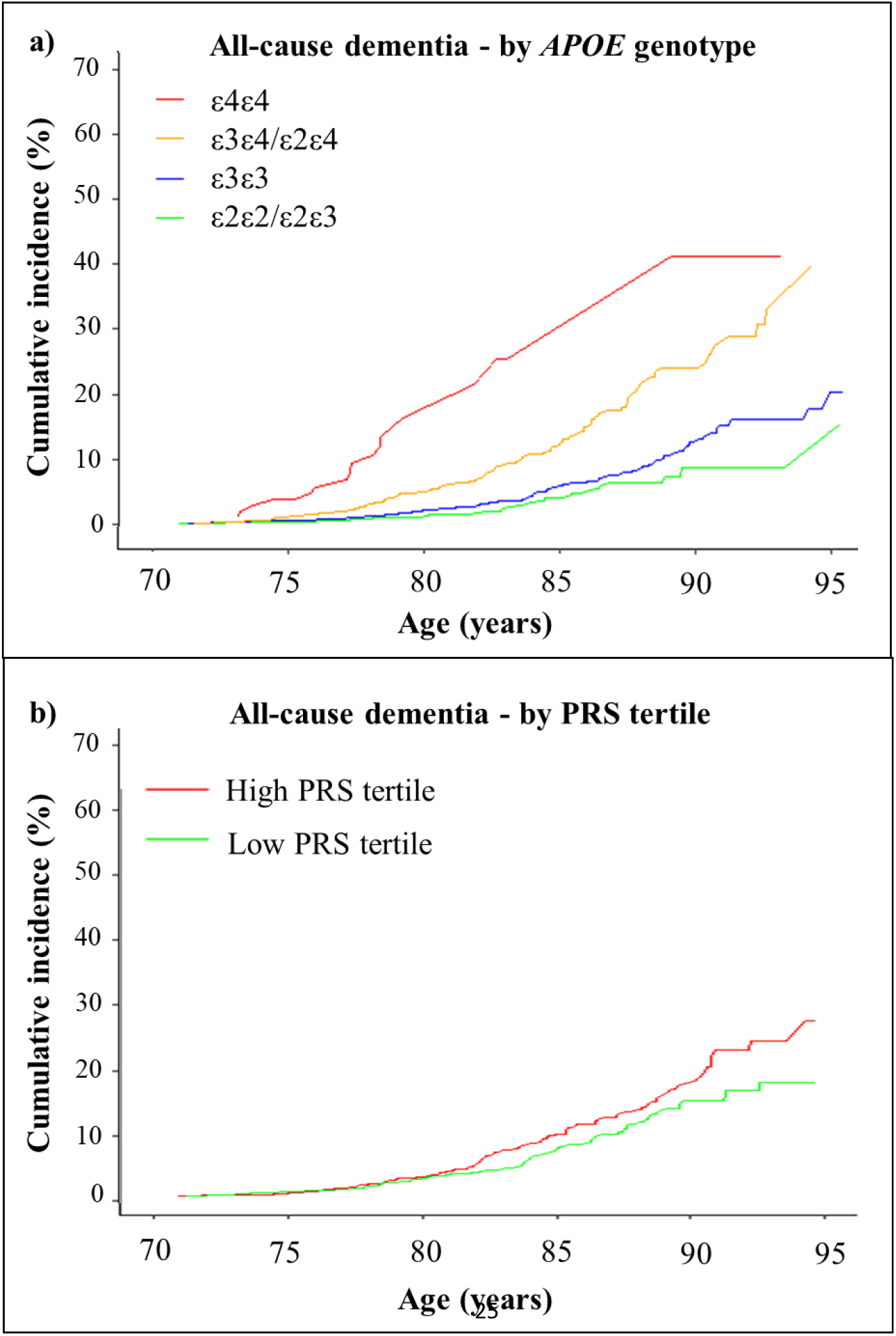
Cumulative incidence of all-cause dementia stratified by *APOE* genotypes and tertiles of a polygenic risk score (PRS). Cumulative incidence curves for all-cause dementia (**a**) and cognitive decline (**b**) were calculated to age 95 years and stratified by *APOE* genotype, with mortality as a competing event. Confidence intervals and participants at risk are shown in Table S3-4.

Dementia risk was higher for participants in the high-risk PRS tertile than the low (HR 1.36 [CI 1.0- 1.7], P=0.02)(Table 3a). CID in the high-risk tertile was 9.6% (CI 7.8-11.8) compared with the low tertile 7.3% (CI 5.7-9.3)(Figure 1b; Table S4). At age 95 years, the effect of PRS was more prominent with CID increasing from 17.6% (13.4-23.0) in the low PRS tertile to 30.6% (21.9-41.9) in the high (Table S4).

In sub-group analysis, among *APOE* ε3/ε4 heterozygotes, PRS modified dementia risk, with CID increasing from 10.8% (CI 7.2-16.3) in the low PRS tertile to 17.8% (CI 13.2-23.8) in the high (Table S5). Among *APOE* ε4/ε4 homozygotes with high PRS (the highest genetic risk category), CID was 32.2% (CI 11.3-71.6). In ε4/ε4 homozygotes with low PRS, CID was lower at 24.6% (CI 11.2-48.8). For ε3/ε3 homozygotes, CID in the low-risk PRS tertile was 5.7% (CI 3.9-8.3) and in the high-risk 7.6% (CI 5.5-10.5)(Table S5).

We compared CID between the highest genetic risk group at age 80 (ε4 carriers with high PRS) and the lowest genetic risk group at age 90 (ε2 carriers with low PRS). CID in the highest genetic risk group at age 80 was 6.1% (CI 4.1-9.0) and in the lowest genetic risk at age 90 was 8.8% (CI 4.5-16.7) (Table S5). This corresponded to an approximately 10-year delay in age-of-onset between these two extreme groups. In sensitivity analysis, we examined interaction between *APOE/*PRS, and found no significant association with incident dementia (P>0.05).

A total of 1598 (12.6%) participants had cognitive decline (Table 1). The cumulative incidence of cognitive decline to age 85 years was estimated to be 37.2% (CI 36.4-41.0) in *APOE* ε3/ε3 homozygotes, 35.3% (CI 30.5-39.6) in ε2/ε2 homozygotes, 45.7% (CI 46.5-53.9) in ε3/ε4 heterozygotes, and 52.9% (CI 46.1-76.2) in ε4/ε4 homozygotes (Figure 2a, Table S6). Compared with the ε3/ε3 reference group, cognitive decline risk was significantly higher in ε3/ε4 heterozygotes (HR=1.35 [1.20-1.51], P<0.001) and ε4 homozygotes (HR=1.75 [CI 1.24-2.46]), P<0.001)(Table 3b).PRS was not associated with cognitive decline. Risk of cognitive decline did not significantly increase between low and high PRS tertiles (HR=1.08 [0.96-1.22], P=0.18)(Figure 2b, Table S7-S8). In sensitivity analysis, the interaction effect between *APOE*/PRS for cognitive decline was not significant (P>0.05).

**Figure 2:**
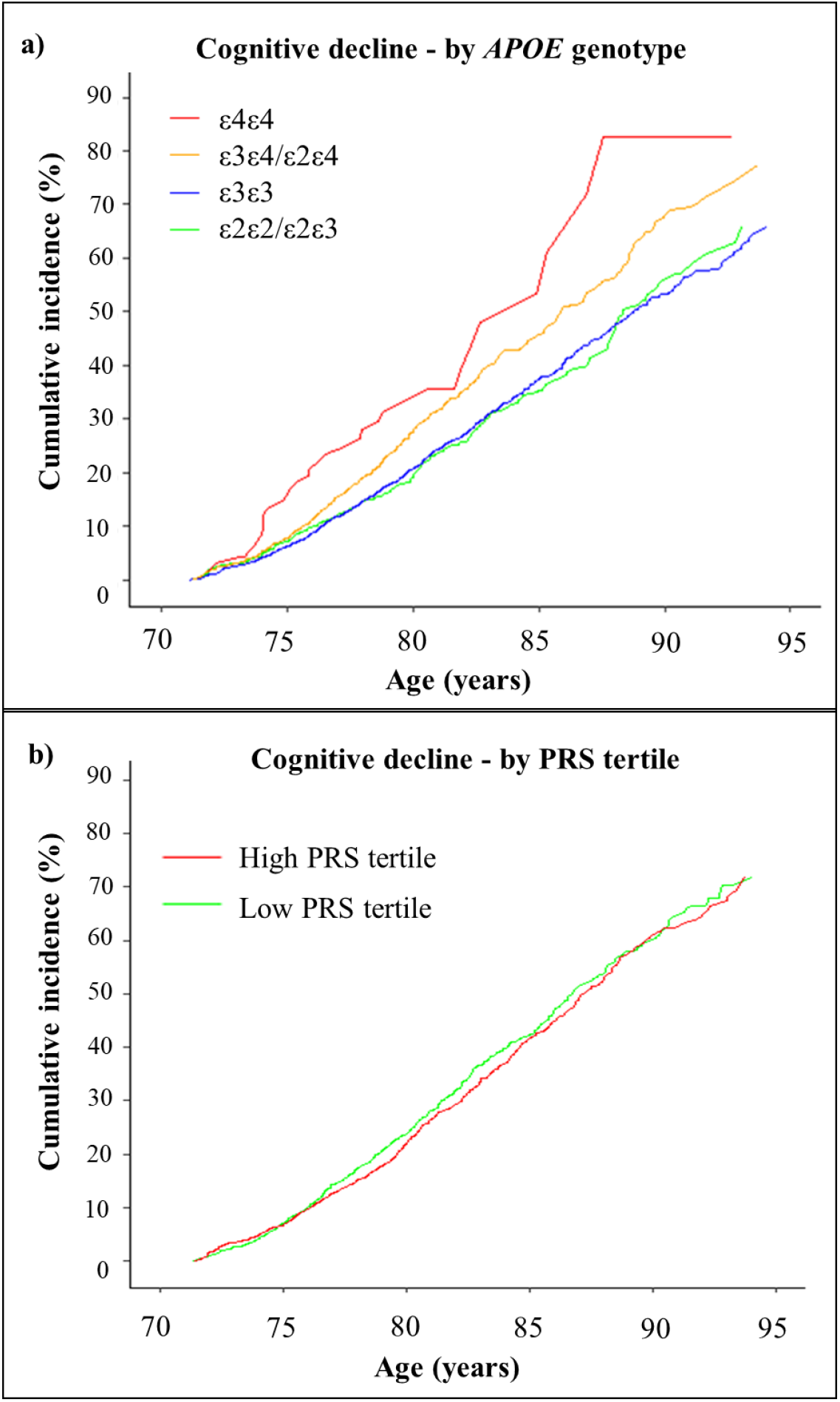
Cumulative incidence of cognitive decline stratified by *APOE* genotypes and tertiles of a polygenic risk score (PRS). Cumulative incidence curves for cognitive decline were calculated with mortality as a competing event, stratified by *APOE* genotypes (**a**) tertiles of a PRS (**b**) based on 23 common non-*APOE* variants. High PRS is shown in red; low PRS in green (mid tertile not shown). Confidence intervals and participants at risk are shown in Table S6-7.

## DISCUSSION

In this study, we examined the effect of *APOE* genotypes and PRS on incident dementia and cognitive decline among 12,978 initially healthy older participants. We found that *APOE* ε4 and high PRS were associated with increased relative risk of dementia, but overall, cumulative incidence of dementia was low across all genotype groups. PRS effect on dementia risk was modest and delayed compared with *APOE* ε4, mostly affecting risk after 85 years of age. *APOE* ε4 was associated with cognitive decline, but PRS was not, suggesting that *APOE* genotype has a stronger effect than PRS on both dementia and cognitive decline. We observe that the absence of co-morbidities, atherothrombotic cardiovascular disease and cognitive impairment to age 70 years contributed to the attenuation of incident dementia across all genotypes.

The unique ascertainment of the ASPREE population is an important factor in the interpretation of our results. The eligibility criteria excluded individuals with dementia diagnoses and cognitive impairment at enrollment, and individuals with any history or diagnosis of atherothrombotic cardiovascular disease events, major physical disability or life-threatening cancer(1). This produced a highly selected population of healthy older individuals, who at the time of study entry, benefited from the absence of several important dementia risk factors. This selective pressure resulted in deviation from the Hardy-Weinberg equilibrium, with fewer deleterious *APOE* ε4 alleles observed than expected. Selection against ε4 was driven by the age cut-off (>70 years), as well as the strict inclusion/exclusion criteria of the trial.

We accordingly observed a low cumulative incidence of all-cause dementia, estimated to be 7.4% to age 85 across all participants. This estimate was approximately half that reported in the community- based Rotterdam study to the same age (15.6%)(9). While acknowledging potential issues with comparing dementia risk between different studies, including differences in population demography, recruitment criteria, diagnostic definitions, and duration of follow-up(7, 9, 24), we consider comparisons between ASPREE and the Rotterdam study warranted. The studies had similar sample sizes, age ranges, sex percentages, diabetes, hypertension, BMI, blood lipids, genetic ancestry and adjudicated dementia cases. Further, both analyses used the same PRS calculations(17, 24, 36).

The lower risk of dementia in ASPREE is likely influenced by the selection of healthy participants, depletion of deleterious *APOE* ε4 alleles, and a relatively short follow-up period where healthy selection effects have not yet dissipated. The estimated CID in ASPREE was 26.6% for ε4/ε4 homozygotes, compared with approximately 60% in the Rotterdam study, and 5.9% for ε3/ε4 heterozygotes, compared with approximately 25% in the Rotterdam study(9). These differences in CID are substantial, unlikely to be attributable to confounding factors alone between the studies.

Further, in a recent meta-analysis of three population-based cohorts of cognitively normal subjects aged 60-75 years (total N=11,771), the risk of dementia in *APOE* ε4/ε4 homozygotes (N=134) to age 70-75 years was 11.2%(7). In ASPREE, however, the risk of dementia to age 75 in ε4/ε4 (N=200) was only 3.7%. Risk of dementia among ε4/ε4 homozygotes to age 85 years in the Framingham Heart Study (37.6%, N=67) was also considerably higher than ASPREE (26.6% N=200)(7).

PRS is more challenging to interpret across studies, given the different PRSs used(7, 9, 21-24). However, we also observed an attenuated effect of PRS on dementia in ASPREE compared with other studies. We observed only a 2.6% difference in CID between low (7.3%) and high (9.6%) PRS tertiles. In the Rotterdam study, the observed difference was 9.0% between low (11.6%) and high (20.4%) tertiles to the same age.

In ASPREE, the effect of PRS was more pronounced in *APOE* ε4 carriers, compared with the reference ε3/ε3 group. However, the PRS effect was attenuated and delayed in age-of-onset compared with other studies(9, 40). The PRS effect on dementia risk in ASPREE mostly occurred after the age of 85 years (Figure 2a). We found no significant interaction effect between *APOE* and PRS in ASPREE, unlike the Rotterdam study(9). This again may reflect the attenuation of genetic effects on dementia risk in ASPREE. A recent analysis of the Framingham cohort also reported no significant interaction between *APOE* and PRS while evaluating dementia risk(40). Further studies with large populations and longer follow-up are required to understand interactions between *APOE* and PRS in modifying dementia risk.

Considering the attenuated genetic risk of dementia observed in ASPREE, we query whether other factors further modified risk, beyond the low vascular risk, cognitive screening and absence of cardiovascular disease at baseline. Such factors could include a favourable lifestyle, characterized by healthy diet, regular exercise and high socialisation levels(29, 30). Alternatively, the attenuation could be related to the relatively short follow-up period, during which healthy selection effects had not yet dissipated.

Protective genetic loci not included in the PRS may also have contributed to risk modification, including common variants yet to be identified by GWAS, and/or rare high-effect protective variants, including loss-of-function variants in biologically associated genes. There is growing evidence that protection from dementia risk can be conferred by both common and rare genetic variants, especially in the high-risk *APOE* ε4/ε4 group(41, 42). Further studies are required to examine the effect of protective genetic variants for dementia in ASPREE.

*APOE* ε4 carrier status was significantly associated with cognitive decline in ASPREE, but PRS was not. This reflects the more modest effect of PRS on cognitive aging, and/or a divergent genetic aetiology versus *APOE* genotype(26). The association between *APOE* ε4 and cognitive decline in non-demented individuals has been reported by several studies using comparable cognitive testing(10-13, 28). However, few studies have reported a significant effect of PRS on cognitive decline alone(25-28). It appears that PRS derived from GWAS of diagnosed dementia/AD cases are not strong predictors of cognitive decline without dementia during aging. However, our approach to quantifying cognitive decline may be insensitive, or might reflect a different biological process. Alternatively, PRS derived from a GWAS of dementia/AD cases may reflect the functional impairment required for dementia diagnosis, rather than the cognitive aspects.

Strengths of the study include a well-characterized longitudinal cohort with repeated cognitive assessments and dementia adjudication, genetic data for both *APOE* and PRS variants, longitudinal follow-up to enable survival analysis for dementia and cognition, data available on covariates, adjudicated reports of causes of death to control for competing events, and a large number of initially healthy elderly participants.

Limitations of the study include a shorter duration of follow-up compared with other studies(7, 9) (possibly insufficient to overcome a healthy volunteer effect) and limited event numbers in some rarer *APOE* genotype groups. Dementia events were not stratified into AD versus other dementia types. However, very few dementia cases were classified as non-AD in ASPREE (1.2%), with the majority of dementia classified as probable/possible-AD(5).

In conclusion, our study found that *APOE* genotypes and PRS effect the relative risk of dementia in a population of healthy older individuals followed prospectively. However, overall CID in the population was low across all genotype groups, reflecting the healthy nature of the population at enrollment. *APOE* ε4 had a stronger effect than PRS on dementia risk. *APOE* genotypes affected cognitive decline, whereas PRS did not. Prospective studies of initially healthy older participants with longer follow-up periods are required to further understand the genetic risk of dementia and cognitive decline during aging, and examine the predictive performance and clinical utility of PRS.

## Supporting information

Supplement Details

## Data Availability

Contact senior author of the paper for data availability

## ABBREVIATIONS

AD: Alzheimer’s disease
APOE: Apolipoprotein E
ASPREE: ASPirin in Reducing Events in the Elderly
GWAS: Genome-wide association study
PRS: Polygenic risk score
HWE: Hardy-Weinberg equilibrium
CID: Cumulative incidence of dementia

## Acknowledgements

We thank the ASPREE trial staff in Australia and the United States, the ASPREE participants who volunteered for the trial, and the general practitioners and staff of the medical clinics who cared for the participants.

## Funding

The ASPREE Healthy Ageing Biobank was supported by a Flagship cluster (including the Commonwealth Scientific and Industrial Research Organisation, Monash University, Menzies Research Institute, Australian National University, University of Melbourne), Monash University and the National Cancer Institute at the National Institutes of Health (supplement to U01AG029824). The ASPREE clinical trial was supported by the National Institute on Aging and the National Cancer Institute at the National Institutes of Health (U01AG029824). the National Health and Medical Research Council (NHMRC) of Australia (334047 and 1127060), Monash University and the Victorian Cancer Agency. We thank the trial staff in Australia and the US, the participants who volunteered for this trial, and the general practitioners and staff of the medical clinics who cared for the participants. We also thank the UK Biobank participants and admin staff. P.L is supported by a National Heart Foundation Future Leader Fellowship (102604).

## Disclosures

Dr. Shah serves as a non-compensated member of the Board of Directors of the Alzheimer’s Association – Illinois Chapter. His institution, Rush University Medical Center, receives funds for his role as Site Principal Investigator or Site Sub-Investigator for industry sponsored clinical trials and research studies involving Alzheimer’s disease and dementia sponsored by Amylyx Pharmaceuticals, Inc., Eli Lilly & Co., Inc., Genentech, Inc., Merck & Co., Inc., Navidea Biopharmaceuticals, and Novartis Pharmaceuticals, Inc. Dr Sebra serves as Vice-President of Technology Development at Sema4. Dr Schadt serves as Chief Executive Officer at Sema4. Dr Goate has consulted for Biogen AbbVie, GSK, Eisai and Denali Therapeutics. No other conflicts were reported.

## Notes

### Competing Interest Statement

Dr. Shah serves as a non-compensated member of the Board of Directors of the Alzheimers Association Illinois Chapter. His institution Rush University Medical Center receives funds for his role as Site Principal Investigator or Site Sub-Investigator for industry sponsored clinical trials and research studies involving Alzheimers disease and dementia sponsored by Amylyx Pharmaceuticals Inc. Eli Lilly & Co. Inc. Genentech Inc. Merck & Co. Inc. Navidea Biopharmaceuticals and Novartis Pharmaceutical Inc. Dr Sebra serves as Vice President of Technology Development at Sema4. Dr Schadt serves as Chief Executive Officer at Sema4. Dr Goate has consulted for Biogen AbbVie GSK Eisai and Denali Therapeutics. No other conflicts were reported.

### Author Declarations

Informed consent for genetic analysis was obtained with ethical approval from the Alfred Hospital Human Research Ethics Committee (390/15) and site-specific Institutional Review Boards (US).

