## Supplement Details for "Effect of APOE and a polygenic risk score on incident dementia and cognitive decline in a healthy older population"

**Supplementary Material**

### Genotyping and Princpal Component analysis

We followed best practice genotyping and quality control (QC) protocols from Thermo Fisher, starting from raw intensity CEL files, we used a custom script designed for the the AXIOM PMDA array uing the human genome reference GRCh38 , to produce variant call files. We performed sample level QC using plink version 1.9, excluding samples for gender discordance (80 samples mismatched and excluded) using plink default F statistics threshold (≤ 0.2 female and ≥ 0.8 male), relatedness (124 indviduals excluded) using default PI-HAT threshold >0.025 to exclude one sample from each related pair. To estimate population structure in the ASPREE cohort we performed principal component analysis (PCA) using The 1000 Genomes Project as a reference population (24). Directly genotyped data from ASPREE and The 1000 Genomes Project 1K phase 3 (liftover to GRCh38) were merged and LD pruned ($r^2 < 0.1$) using plink version 1.9 (27), followed by R package SNPrelate (25). We calculated the Z score for first 2 principal component eigenvectors and excluded samples with ± 2SD (standard deviation) of Z score compared to their respective five reference superpopulation groups from the 1000 Genomes Project that included: Europenas, South Asians, East Asians, African American (African super population) and Hispanics (Ad Mixed American) (Figure S1). The final dataset of 12,978 Caucasians (Non-Finish Europeans) were selected from the ASPREE cohort for further analysis.

**Figure S1: Principal component analysis (PCA) of the ASPREE cohort compared with the 1000 Genomes Project. A)** PCA plot of all ASPREE participants projected onto 1000 Genome populations. **B).** PCA plot of ASPREE Europeans samples projected onto 1000 Genome Europeans samples that were included in this study. In Figure legend 1K genome populations are (Europeans, South Asians, East Asians, African American and Hispanics. ASPREE_AA is African American samples.

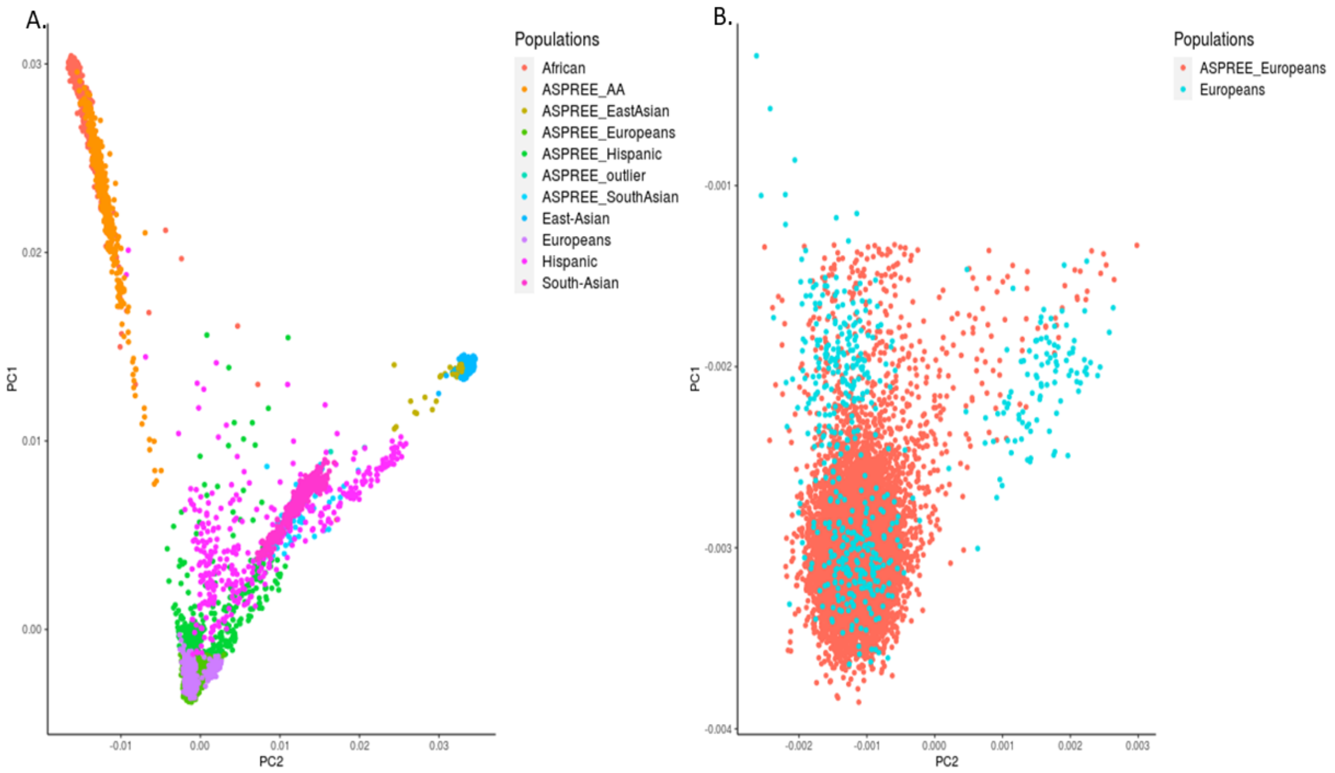

**Table S1: SNPs used in polygenic risk score (PRS) of dementia**

| Gene (NCBI) | SNP | Effect allele | Effect Size (Beta)* | Reference No |
|---|---|---|---|---|
| ABCA7 | rs4147929 | G | -0·135 | 4 |
| BIN1 | rs6733839 | T | 0·188 | 4 |
| CASS4 | rs7274581 | C | -0·139 | 4 |
| CD2AP | rs10948363 | G | 0·098 | 4 |
| CELF1 | rs10838725 | C | 0·075 | 4 |
| CLU | rs9331896 | T | 0·146 | 4 |
| CR1 | rs6656401 | G | -0·157 | 4 |
| ECHDC3 | rs7920721 | G | -0·067 | 20 |
| EPHA1 | rs11771145 | A | -0·102 | 4 |
| FERMT2 | rs17125944 | C | 0·122 | 4 |
| HLA-DRB1/5 | rs111418223 merged into rs9271192 | A | -0·108 | 4 |
| HS3ST1 | rs13113697 | G | -0·067 | 20 |
| INPP5D | rs35349669 | T | 0·066 | 4 |
| KANSL1 | rs118172952 merged into rs2732703 | G | -0·151 | 4 |
| MEF2C | rs190982 | A | 0·08 | 4 |
| MS4A6A | rs983392 | G | -0·108 | 4 |
| NME8 | rs2718058 | G | -0·07 | 4 |
| PICALM | rs10792832 | G | 0·13 | 4 |
| PTK2B | rs28834970 | C | 0·096 | 4 |
| SLC24A4-RIN3 | rs10498633 | T | -0·104 | 4 |
| SORL1 | rs11218343 | C | -0·27 | 4 |
| TREM2 | rs75932628 | T | 0·889 | 21 |
| ZCWPW1 | rs1476679 | T | 0·078 | 4 |

*Shows the weighted effect size used by *Lee et al 2018* (9) to calculate the PRS.

**Table S2. Hardy-Weinberg equilibrium (HWE) testing for *APOE* genotype frequencies in the ASPREE population.** The chi square test was performed to calculate deviation from Hardy-Weinberg equilibrium [Preacher, K. J. (2001, April). Calculation for the chi-square test: An interactive calculation tool for chi-square tests of goodness of fit and independence [Computer software]. Available from http://quantpsy.org.]

| **Alleles: P** = 0.77 (*APOE* ε3), **Q** = 0.13 (*APOE* ε4), **R** = 0.09 (*APOE* ε2) | | | | |
|---|---|---|---|---|
| | **Observed** | **Expected** | **Chi Square** | **P value** |
| ε3ε3 | 7800 | 7743 | 38.0 | P < 0.001 |
| ε3ε4 | 2665 | 2723 | | |
| ε2ε2 | 68 | 109 | | |
| ε2ε3 | 1784 | 1839 | | |
| ε2ε4 | 461 | 323 | | |
| ε4ε4 | 200 | 239 | | |

**Figure S2: Polygenic risk score (PRS) distribution in the ASPREE cohort.** The red dotted line shows boundaries of PRS tertiles; lower risk tertile -0.56 (range: -1.43 to -0.34), middle risk tertile -0.20 (range -0.34 to -0.06) and high risk tertile 0.16 (range -0.06 to 1.86).

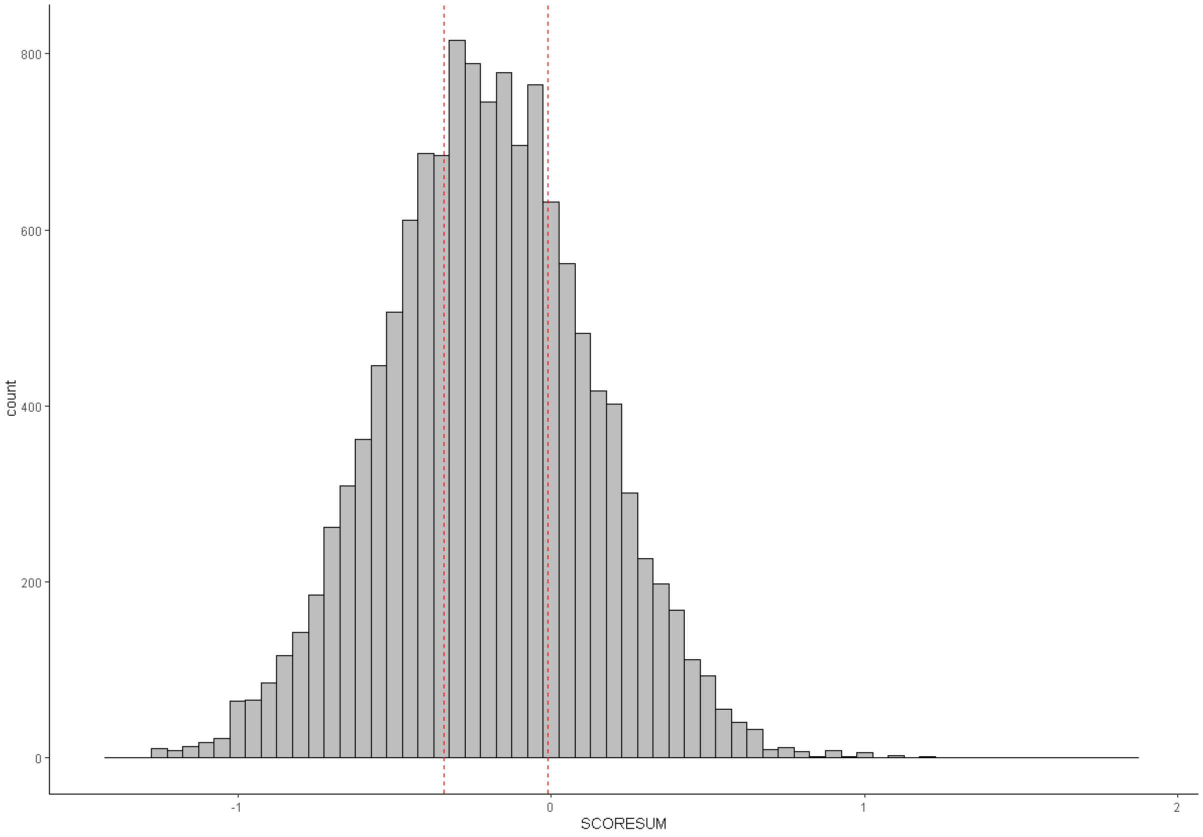

**Table S3: Cumulative incidence of dementia stratified by *APOE* genotypes, accounting for death as a competing risk**

| *APOE* genotypes | Age, years* | Participants at risk N | Risk, [95% Confidence Interval] |
|---|---|---|---|
| ε2/ε2:ε2/ε3 | 75 | 835 | 0.2% [0.0-1.1] |
| ε3/ε3 | 75 | 3672 | 0.4% [0.2-0.7] |
| ε2/ε4:ε3/ε4 | 75 | 1555 | 1.0% [0.6-1.6] |
| ε4/ε4 | 75 | 103 | 3.7% [1.4-9.5] |
| ε2/ε2:ε2/ε3 | 80 | 462 | 1.2% [0.6-2.4] |
| ε3/ε3 | 80 | 1838 | 2.0% [1.5-2.6] |
| ε2/ε4:ε3/ε4 | 80 | 719 | 4.9% [3.8-6.3] |
| ε4/ε4 | 80 | 40 | 19.3% [11.9–30.5] |
| ε2/ε2:ε2/ε3 | 85 | 217 | 4.0% [2.4-6.5] |
| ε3/ε3 | 85 | 762 | 5.9% [4.8-7.2] |
| ε2/ε4:ε3/ε4 | 85 | 258 | 12.6% [10.2-15.5] |
| ε4/ε4 | 85 | 17 | 26.6% [16.2–42.0] |
| ε2/ε2:ε2/ε3 | 90 | 38 | 8.7% [5.4-14.1] |
| ε3/ε3 | 90 | 193 | 5.9% [4.8-7.2] |
| ε2/ε4:ε3/ε4 | 90 | 58 | 12.6% [10.2-15.5] |
| ε4/ε4 | 90 | 3 | 26.6% [16.2–42.0] |
| ε2/ε2:ε2/ε3 | 95 | 7 | 15.4% [6.5-34.0] |
| ε3/ε3 | 95 | 17 | 21.2% [15.0-2.5] |
| ε2/ε4:ε3/ε4 | 95 | 5 | 42.0% [28.7-58.4] |
| ε4/ε4 | 95 | 1 | 48.5% [20.5–85.3] |

*Five-year longitudinal age from baseline to dementia or censored

**Table S4: Cumulative incidence of dementia stratified by tertiles of a polygenic risk score (PRS), accounting for death as a competing risk**

| PRS Tertiles | Age, years | Participants at risk N | Risk, [95% Confidence Interval] |
|---|---|---|---|
| Low | 75 | 2038 | 0.6% [0.3–1.1] |
| Middle | 75 | 2024 | 0.7% [0.4-1.2] |
| high | 75 | 2096 | 0.4% [0.2-0.8] |
| Low | 80 | 1020 | 2.7% [2.0–3.6] |
| Middle | 80 | 1030 | 3.0% [2.2-4.0] |
| high | 80 | 1021 | 2.9% [2.2-3.9] |
| Low | 85 | 410 | 7.3% [5.7–9.3] |
| Middle | 85 | 429 | 5.8% [4.4-7.5] |
| high | 85 | 411 | 9.6% [7.8-11.8] |
| Low | 90 | 116 | 14.7% [11.5–18.6] |
| Middle | 90 | 88 | 15.3% [11.6-19.9] |
| high | 90 | 9 | 17.6% [14.2-21.8] |
| Low | 95 | 12 | 17.6% [13.4–23.0] |
| Middle | 95 | 8 | 27.8% [16.9-43.4] |
| high | 95 | 9 | 30.6% [21.9-41.9] |

*Five-year longitudinal age from baseline to dementia or censored

### Table S5: Cumulative incidence of dementia stratified by *APOE* genotypes and tertiles of a PRS

| | | APOE:e3e3 | | APOE:e3e4ande2e4 | | APOE:e2e2&e2e3 | | APOE:e4e4 | |
|---|---|---|---|---|---|---|---|---|---|
| **PRS Group** | **Age** | **Risk, 95% CI** | **N** | **Risk, 95% CI** | **N** | **Risk, 95% CI** | **N** | **Risk, 95% CI** | **N** |
| Low tertile | 75 | 0.6%, 0.2 – 1.2 | 1200 | 0.6%, 0.2 – 1.9 | 534 | 0.9%, 0.2 – 3.5 | 270 | 5.4%, 1.4 – 20.0 | 36 |
| Middle tertile | 75 | 0.3%, 0.1 – 1.0 | 1197 | 1.5%, 0.7 – 3.1 | 513 | 0.0%, 0.0 -0.0 | 282 | 6.1%, 1.6 – 22.5 | 36 |
| High tertile | 75 | 0.2%, 0.1 – 0.9 | 1277 | 1.0%, 0.4 – 2.4 | 505 | 0.0%, 0.0 -0.0 | 283 | 0.0%, 0.0 – 0.0 | 33 |
| Low tertile | 80 | 2.0%, 1.3 – 3.1 | 606 | 4.1%, 2.5 – 6.8 | 237 | 2.1%, 0.8 – 5.5 | 147 | 18.2%, 7.7 – 39.7 | 15 |
| Middle tertile | 80 | 2.3%, 1.5 – 3.6 | 608 | 4.6%, 3.0 – 7.1 | 251 | 0.4%,0.1 -2.8 | 144 | 23.6%, 11.9 – 43.4 | 22 |
| High tertile | 80 | 1.8%, 1.1 – 2.9 | 595 | 6.1%, 4.1 – 9.0 | 237 | 1.2%, 0.4 – 3.6 | 166 | 16.6%, 5.3 – 45.3 | 12 |
| Low tertile | 85 | 5.7%, 3.9 – 8.3 | 260 | 10.8%, 7.2 – 16.3 | 77 | 7.1%, 3.6 – 13.7 | 66 | 24.6%, 11.2 – 48.8 | 9 |
| Middle tertile | 85 | 4.8%, 3.2 – 7.0 | 266 | 9.1%, 6.0 – 13.8 | 95 | 3.4%, 1.2 – 9.3 | 68 | | |
| High tertile | 85 | 7.6%, 5.5 – 10.5 | 240 | 17.8%, 13.2 – 23.8 | 90 | 2.9%, 1.1 – 7.4 | 77 | 32.2%, 11.3 -71.6 | 5 |
| Low tertile | 90 | 11.6%, 8.1 – 16.6 | 80 | 24.4%, 16.9 – 34.5 | 28 | 8.8%, 4.5 – 16.7 | 17 | | |
| Middle tertile | 90 | 14.4%, 9.6 – 21.5 | 53 | 21.6%, 14.8 – 30.9 | 22 | 7.9%, 3.2 – 18.8 | 6 | | |
| High tertile | 90 | 15.2%, 11.1 – 20.6 | 67 | 32.3%, 22.8 – 44.3 | 15 | 9.6%, 3.8 – 23.1 | 16 | | |
| Low tertile | 95 | 14.0%, 9.6 – 20.2 | 8 | 31.9%, 19.0 – 50.4 | 6 | 8.8%, 4.5 – 16.7 | 6 | | |
| Middle tertile | 95 | 28.5%, 14.5 – 51.2 | 6 | 32.3%, 21.1 – 47.3 | 8 | 7.9%, 3.2 – 18.8 | 2 | | |
| High tertile | 95 | 21.3%, 15.4 – 29.3 | 7 | 76.8%, 61.4 – 89.4 | 1 | 19.5%, 7.0 – 47.7 | 4 | | |

APOE: Apolipoprotein E, Dementia risk is determine using 5 years of age intervals accounted for death as competing risk, The cumulative incidence was calculated up to 95 years of age, beyond age 95 the data was very limited with no participant alive or demented also data was also sparse, similar case for APOE e4e4 beyond age 85 . N is number of participants at risk is for APOE categories for each PRS tertile. Empty cells show data was sparse or that the participants at those ages and APOE/PRS categories were not demented or alive.

**Table S6: Cumulative incidence of cognitive decline stratified by *APOE* genotypes, accounting for death as a competing risk**

| *APOE* genotypes | Age, years* | Participants at risk N | Risk, [95% Confidence Interval] |
|---|---|---|---|
| ε2/ε2:ε2/ε3 | 75 | 668 | 7.3% [5.4-9.8] |
| ε3/ε3 | 75 | 2966 | 6.2% [5.4-7.2] |
| ε2/ε4:ε3/ε4 | 75 | 1233 | 7.8% [6.3-9.5] |
| ε4/ε4 | 75 | 76 | 15.9 %[9.7-25.4] |
| ε2/ε2:ε2/ε3 | 80 | 352 | 19.0% [16.1-22.4] |
| ε3/ε3 | 80 | 1346 | 20.8% [19.2-22.4] |
| ε2/ε4:ε3/ε4 | 80 | 504 | 27.5% [24.9-30.4] |
| ε4/ε4 | 80 | 34 | 33.1% [23.6–45.1] |
| ε2/ε2:ε2/ε3 | 85 | 155 | 35.3% [30.5-39.6] |
| ε3/ε3 | 85 | 553 | 37.2% [36.4-41.0] |
| ε2/ε4:ε3/ε4 | 85 | 174 | 45.7% [46.5-53.9] |
| ε4/ε4 | 85 | 9 | 52.9% [46.1–76.2] |
| ε2/ε2:ε2/ε3 | 90 | 26 | 54.9% [48.4-61.7] |
| ε3/ε3 | 90 | 134 | 52.9% [49.7-56.1] |
| ε2/ε4:ε3/ε4 | 90 | 33 | 67.3% [62.0-72.4] |
| ε4/ε4 | 90 | 2 | 81.8% [59.6-95.9] |
| ε2/ε2:ε2/ε3 | 95 | 6 | 63.7% [55.2-72.1] |
| ε3/ε3 | 95 | 12 | 64.4% [59.8 -68.9] |
| ε2/ε4:ε3/ε4 | 95 | 4 | 75.1% [68.0-81.7] |
| ε4/ε4 | 95 | 1 | 81.8% [59.6-95.9] |

*Five-year longitudinal age from baseline to cognitive decline or censored

**Table S7: Cumulative incidence of cognitive decline stratified by tertiles of a polygenic risk score (PRS), accounting for death as a competing risk**

| PRS Tertiles | Age, years* | Participants at risk N | Risk, [95% Confidence Interval] |
|---|---|---|---|
| Low | 75 | 1662 | 6.7% [5.5-8.1] |
| Middle | 75 | 1608 | 6.8% [5.6-8.2] |
| high | 75 | 1677 | 7.3% [6.1-8.7] |
| Low | 80 | 748 | 21.5% [19.4-23.8] |
| Middle | 80 | 752 | 22.7% [20.5-25.0] |
| high | 80 | 730 | 22.9% [20.8-25.2] |
| Low | 85 | 283 | 38.9% [35.8-42.2] |
| Middle | 85 | 315 | 38.7% [35.7-41.9] |
| high | 85 | 290 | 39.8% [36.7-43.0] |
| Low | 90 | 75 | 57.0% [52.7-61.4] |
| Middle | 90 | 60 | 58.2% [53.7-62.8] |
| high | 90 | 65 | 55.5% [51.4-59.8] |
| Low | 95 | 4 | 66.3% [60.5-72.0] |
| Middle | 95 | 2 | 74.5% [63.9-83.9] |
| high | 95 | 6 | 66.8% [61.0-72.4] |

*Five-year longitudinal age from baseline to cognitive decline or censored

### Table S8: Cumulative incidence of cognitive decline stratified by *APOE* genotypes and tertiles of a PRS

| | | APOE:e3e3 | | APOE:e3e4ande2e4 | | APOE:e2e2&e2e3 | | APOE:e4e4 | |
|---|---|---|---|---|---|---|---|---|---|
| **PRS Group** | **Age** | **Risk, 95% CI** | **N** | **Risk, 95% CI** | **N** | **Risk, 95% CI** | **N** | **Risk, 95% CI** | **N** |
| Low tertile | 75 | 5.9%, 4.5 – 7.7 | 987 | 6.3%, 4.3 – 9.3 | 422 | 8.9%, 5.4 – 14.5 | 223 | 27.5%, 14.6 – 48.1 | 27 |
| Middle tertile | 75 | 6.2%, 4.8 – 8.1 | 961 | 7.8%, 5.4 – 11.1 | 401 | 7.5%, 4.6 – 12.0 | 228 | 15.8%, 6.3 – 36.9 | 24 |
| High tertile | 75 | 6.6%, 5.2 – 8.4 | 1029 | 9.5%, 6.9 – 13.1 | 401 | 6.2%, 3.6 – 10.6 | 223 | 8.6%, 2.8 – 24.3 | 28 |
| Low tertile | 80 | 19.6%, 17.0 – 22.5 | 462 | 25.9%, 21.5 – 31.0 | 169 | 19,4%, 14.3 – 26.0 | 114 | 46.0%, 29.6 – 66.1 | 12 |
| Middle tertile | 80 | 21.1%, 18.5 – 24.1 | 460 | 28.1%, 23.5 – 33.3 | 169 | 20.4%,15.5 – 26.7 | 110 | 31.9%, 15.9 – 57.2 | 10 |
| High tertile | 80 | 21.5%, 18.9 – 24.5 | 431 | 29.4%, 24.8 – 34.5 | 163 | 18.1%, 13.5 – 24.0 | 127 | 49.7%, 19.7 – 88.5 | 3 |
| Low tertile | 85 | 37.8%, 33.8 – 42.1 | 187 | 43.7%, 37.0 – 50.9 | 47 | 35.7%, 28.2 – 44.5 | 44 | 52.0%, 34.1 – 72.6 | 6 |
| Middle tertile | 85 | 36.6%, 32.8 – 40.6 | 194 | 47.0%, 40.8 – 53.6 | 66 | 33.2%, 26.1 – 41.6 | 54 | 72.7%, 34.0 – 98.3 | 2 |
| High tertile | 85 | 37.6%, 33.6 – 41.8 | 171 | 46.6%, 40.3 – 53.3 | 64 | 38.1%, 31.0 – 46.4 | 54 | 73.2%, 35.9 – 98.0 | 2 |
| Low tertile | 90 | 55.8%, 50.4 – 61.4 | 53 | 63.4%, 54.1 – 72.7 | 11 | 55.0%, 44.0 – 67.2 | 13 | 52.0%, 34.2 – 72.6 | 6 |
| Middle tertile | 90 | 52.7%, 47.1 – 58.6 | 39 | 71.5%, 62.8 – 79.8 | 13 | 53.6%, 42.8 – 65.1 | 5 | | |
| High tertile | 90 | 50.9%, 45.7 – 56.4 | 49 | 68.8%, 59.5 – 77.7 | 10 | 55.0%,44.3 – 65.8 | 9 | 96.7%, 85.7 – 99.8 | 1 |
| Low tertile | 95 | 66.0%, 58.4 – 73.5 | 3 | 65.6%, 56.1 – 75.0 | 2 | 66.1%, 52.7 – 79.1 | 4 | | |
| Middle tertile | 95 | 80.0%, 73.4 – 85.9 | 1 | 79.6%, 68.2 – 89.0 | 2 | 53.6%, 42.8 – 65.1 | 4 | | |
| High tertile | 95 | 63.1%, 55.4 – 70.8 | 5 | 77.7%, 65.8 – 87.8 | 2 | 63.4%, 50.4 – 76.2 | 4 | | |

APOE: Apolipoprotein E, Cognitive decline incidence is measured using 5 years of age intervals accounted for death as a competing risk. The cumulative incidence was calculated up to 95 years of age, beyond age 95 the data was limited N is the number of participants at risk APOE categories for each PRS tertile, Empty cells show data was sparse or that the participants at those ages and APOE/PRS categories were not cognitive declined or alive.
